# Integrative genomics elucidates the evolutionary, temporal, and developmental origins of a hydrocephalus risk gene

**DOI:** 10.1101/2025.09.01.25334358

**Authors:** Andrew T. Hale, Yuwei Song, Caroline Davies, Shanrun Liu, Regan Gaskin, Anastasia Arynchyna-Smith, Brandon G. Rocque, Zechen Chong

## Abstract

**Introduction:** A prior integrative, multi-omics human genetics and functional genomics study identified maelstrom (*MAEL*), a gene involved in regulation of DNA transposon activity and genome structure, as a transcriptome-wide predictor of hydrocephalus (HC) in the brain cortex. Here we expand on this discovery and further characterize the evolutionary origin and expression of *MAEL* across developmental timescales and cell-lineages in the neonatal human brain towards a mechanistic understanding how variation in *MAEL* expression may cause HC.

**Objective:** To characterize the evolutionary, temporal, developmental, and lineages of *MAEL* expression in HC and the developing human brain.

**Methods:** Ensembl was used to delineate the evolution and taxonomy of *MAEL* across species. Analysis of single-cell RNA sequencing (scRNA-seq) of 49 brain regions across pre- and post-natal timescales from the Developing Human Brain Atlas (Allen Institute) identified temporal and spatial *MAEL* expression patterns. We quantified *MAEL* expression in primary cortical brain tissue obtained during the surgical treatment of HC.

**Results:** We performed taxonomic gene-mapping to define the evolutionary origin of *MAEL* to assess suitability for mechanistic characterization *in vitro* and *in vivo* across species. We find that *MAEL* is among the top 0.01% human-specific genes and < 50% sequence homology among commonly used model organisms with highly divergent functions, necessitating mechanistic validation in human tissue. scRNA-seq of the non-disease prenatal human brain identified *MAEL* expression enriched in cortical excitatory neurons, which was recapitulated in primary HC brain tissue obtained during surgery. Finally, using scRNA-seq of primary HC brain tissue, we functionally validated reduced *MAEL* expression, consistent with a prior human TWAS analysis.

**Conclusions:** We identify the evolutionary, temporal, and developmental expression pattern of *MAEL* in the neonatal human brain. We also provide direct evidence for reduced *MAEL* expression in human HC brain tissue. These data, at least in part, implicate reduced *MAEL* expression underlying human HC across etiologies.

## INTRODUCTION

Hydrocephalus (HC) is a heterogeneous neurodevelopmental disorder resulting from aberrant brain-cerebrospinal fluid (CSF) homeostasis which leads to elevated intracranial pressure ^1–3^. HC is one of the most common indications for neurological surgery in the world, especially in children ^4,5^, but surgical treatment options are prone to failure. The current treatments for HC include insertion of a ventricular CSF shunt and endoscopic third ventriculostomy (ETV) with or without choroid plexus cauterization (CPC) ^1,6^. While clinical trials have attempted pharmacological strategies to treat HC ^7^, these approaches have been unsuccessful, largely due to an incomplete understanding of underlying molecular mechanisms ^2,3,8,9^. Moreover, many studies have evaluated the efficacy and cost of HC surgical interventions ^10^, but long-term morbidity remains unacceptably high. Key obstacles to non-surgical treatments is our incomplete mechanistic and genetic understanding of the disease ^11^. We believe that agnostic human genetic approaches to identify HC genes and mutations coupled with detailed mechanistic understanding can lead to precision surgical and pharmacological treatments.

There are a number of challenges to identifying the genetic etiology for many cases of HC including phenotypic variability, comorbid neurodevelopmental disorders, polygenicity, and pleiotropy. Varying approaches have been used to understand the genetic basis of HC including trio-based whole-exome sequencing (WES) to identify inherited and *de novo* mutations^12,13^ and genome-wide & transcriptome-wide association studies (GWAS/TWAS)^14,15^, where TWAS identifies gene-level associations with the disease of interest. Prior work from our group using multi-omics analysis, including human TWAS, of HC patients of European ancestry identified maelstrom (*MAEL*) in the brain cortex reaching experiment-wide significance as the leading gene-tissue pair associated with HC ^14^. The data provide the premise for furthering our understanding of *MAEL* in human brain development and pathways relevant to HC pathobiology.

Here, we use functional genomics approaches to identify the evolutionary, temporal, and spatial cellular origin of *MAEL* expression in the developing human brain. We perform taxonomic analysis of *MAEL* to identify the evolutionary origin of the gene across species to prioritize model systems for downstream mechanistic studies. Based on the hypothesized role of MAEL in non-brain cell types regulating DNA structure, we estimate the global and single cell copy number variant (CNV) burden across prenatal time in the human brain. Using large-scale single-cell RNA sequencing (scRNA-seq) atlases of the neonatal human brain, we identify the cellular and temporospatial origins of *MAEL* which correlate with cell types enriched for CNV, consistent with the reported role of *MAEL* in non-brain tissues. Finally, we analyze scRNA-seq data from primary human HC brain tissue, recapitulating reduced M*AEL* expression and functionally validating a human HC TWAS analysis^14^.

Intriguingly, these data also identify *MAEL* expression enriched in excitatory neurons (ENs), which may also explain frequently diagnosed comorbid disorders associated with HC, including autism spectrum disorder (ASD), epilepsy, etc. These data illuminate the complex genetic, cellular, and molecular architecture of HC to offer insights into how *MAEL* may contribute to HC pathobiology. Moreover, these analyses highlight the need for a human model system to elucidate human HC specific disease mechanisms.

## METHODS

### Evolutionary gene-mapping and phylogenetic analysis

Taxonomic and gene-tree analysis was performed using Ensemble ^16^. Genome assembly was referenced to the GRCh38 build. Quantification of gene family expansion and contraction was performed using Computation Analysis of gene Family Evolution (CAFÉ) ^17^. Data visualization and comparative mapping were performed through the Ensembl genome browser interface ^16^. Gene tree analysis identified 127 total speciation nodes and 2 duplication events across the evolutionary trajectory of *MAEL* ^18^.

### Single-cell RNA sequencing of neonatal brain development

scRNA-seq data was obtained from atlases of the neonatal human brain ^19,20^ and the Allen Brain Institute Developing Brain Atlas ^21,22^. BrainSpan Developmental Transcriptome data is a broad developmental survey of gene expression containing 13 developmental stages in 26 brain structures^22^. Developmental stages were selected and regrouped into two groups including early prenatal (8-12 post-conception weeks (pcw)), early mid-prenatal (13-18 pcw), late mid-prenatal (19-24 pcw), late prenatal (25-38 pcw), early infancy (Birth-5 month), late infancy (6-18 months), early childhood (19 months – 5 years), late childhood (6-11 years), adolescence (12-19 years) and adulthood (20-60+ years). Late prenatal stage (25-38 pcw) was also subdivided into four subgroups including late prenatal 25 pcw, late prenatal 26 pcw, late prenatal 35 pcw, and late prenatal 37 pcw reflecting critical developmental windows. FastQC was used for quality control ^23^, then reads were mapped to GENCODE human reference genome (GRCh38.p13 Release 43) using STAR v2.7 ^24^. Transcript quantification was done by RSEM ^25^. For BrainSpan data, only normalized expression data was able to be downloaded and that’s what we used for further analysis.

Differential expression analysis was performed using DESeq2 (version 1.38.3) ^26^ on the merged gene counts from RSEM output and BrainSpan data. Differentially expressed genes were selected based on likelihood ratio test, and an adjusted p-value less than 0.05 after apeglm LFC shrinkage ^27^. BiomaRt was used for gene description annotation for differentially expressed gene and converted from Ensembl to Entrez and HGNC symbols (version 2.54.1) ^28,29^. Heatmaps were plotted using rlog normalized counts with hierarchical clustering and average RPKM expression value for individual structure and developmental stage for BrainSpan. Differential expression plots were for *MAEL,* and each brain structure collected from BrainSpan. FASTQ files were processed using the University of Alabama at Birmingham (UAB) Cheaha supercomputer. All other analyses were performed on a 2021 MacBook Pro using R version 4.2.2 RStudio 2022.12.0+353 “Elsbeth Geranium” Release (2022-12-15) for macOS.

### Single-cell RNA sequencing of human hydrocephalic brain tissue

Patients with HC undergoing endoscopic third ventriculostomy (ETV) for treatment of HC were prospectively identified and consented for inclusion in this study. All aspects of this study were approved by the University of Alabama at Birmingham Institutional Review Board. After the endoscope (without any instruments in the working channel and without a sheath) is introduced into the ventricle, the brain ‘core’ tissue is aspirated in cerebrospinal fluid (CSF) and placed in a specimen collection cup prior to flash freezing and storage at -80°C prior to sample preparation for single-cell RNA sequencing (scRNA seq). Brain tissues were lysed with lysis buffer (Sigma catalog# NUC101) containing RNase inhibitor (Sigma, PN-3335399001, 0.2u/ul) to release nuclei from cells. Lysed brain tissues were stained with 7AAD and 7AAD+ nuclei were flow sorted using BD FACSAria flow cytometer. Single nuclei mRNA was barcoded and underwent reverse transcription and library construction according to 10xGenomics 3’ NextGem 3.1 kit instructions (PN-1000268). Constructed Libraries were sequenced using the Illumina Novaseq 6000 machine targeting 40,000 reads/cell with the sequencing cycle consisting of 28bp for read 1, 90bp for read 2 and 10bp for i7 and i5. Data were analyzed using 10xGenomics Cell Ranger. FASTQ files scRNA seq reads were aligned to the human GRCh38 genome, and gene counts were quantified using Cell Ranger ‘count’, as previously described ^30^. Reads mapping to introns and exons were counted according to 10X Genomics instructions. Seurat was used for downstream analysis ^31^. Quality control metrics included at least 25 genes were expressed and 1,000 - 70,000 transcripts were detected per cell. Read counts were normalized and scaled by the NormalizeData and ScaleData functions of Seurat, respectively. All datasets were preprocessed and integrated using the Seurat (v4.0) workflow in R, with quality control filters applied to remove low-quality nuclei and doublets. Differential expression between hydrocephalus and control samples was assessed using Seurat’s FindMarkers function with the MAST test, which accounts for the zero-inflated nature of single-cell data. To specifically evaluate MAEL, expression values were extracted along with cell type, group (CTRL vs HC), and sample identifiers. Cells with zero MAEL expression were excluded, and group-wise comparisons within each cell type were performed using the Wilcoxon rank-sum test. P values were adjusted for multiple testing using the Benjamini–Hochberg method, and statistical significance was reported as *** p < 0.001; ** p < 0.01; * p < 0.05. To determine whether the average log_2_ fold change (log_2_FC) of *MAEL* expression deviated from zero across samples, we performed a one-sample Wilcoxon signed-rank test. For visualization, boxplots were used to display log_2_FC distributions across hydrocephalus samples, as well as MAEL expression across cell types, with outliers suppressed and significance levels annotated above each comparison. Age-matched reference data from Herring et al.^20^ was used as a control.

## RESULTS

### Gene-mapping and taxonomic analysis reveals evolutionary origin of MAEL

Since *MAEL* is one of the most tissue-specific genes across the genome, and *MAEL* was not replicated in mouse models of HC ^14^, we hypothesized that the gene architecture and function of *MAEL* may be poorly conserved across species. To test this, we used Ensemble ^16^ to perform comparative taxonomic analysis and phylogenetic reconstruction of *MAEL* across species (Figure 1). Phylogenetic analysis identified *MAEL* orthologs across a wide range of taxa, with the earliest identifiable homolog detected in *Drosophila melanogaster*, the species in which the gene was initially discovered, cloned, and characterized ^32^. The reconstructed gene tree revealed 127 total speciation nodes and 2 duplication events across the evolutionary trajectory of *MAEL*. Notably, we observe substantial sequence divergence across mammalian lineages. Within primates, sequence homology among great apes, including humans, was reduced to less than 66% (Figure 1). Furthermore, the Gorilla *MAEL* gene, representing a lineage that diverged just prior to the human-chimpanzee split, contains unique 5’ domain features that are not present in any other great ape species (Figure 1). In addition, the chimpanzee *MAEL* homolog, despite the close evolutionary relationship between humans and chimpanzees, displays higher sequence similarity to Old and New World monkey orthologs than to the human gene. This discordance between expected phylogenetic proximity and observed sequence homology underscores the complexity of *MAEL* evolution and highlights why functional and structural characterization of *MAEL* in mammalian species has been challenging ^33–35^. While there is no statistically significant expansion/contraction of *MAEL* copy number across evolutionary time (p=0.95^17^), it is clear that the gene’s structure and sequence is highly variable across species. Evolutionary and comparative genomics analysis of *MAEL*, thus reaffirms the necessity to study the function of the gene in conferring risk to HC in a human model system.

**Figure 1.**
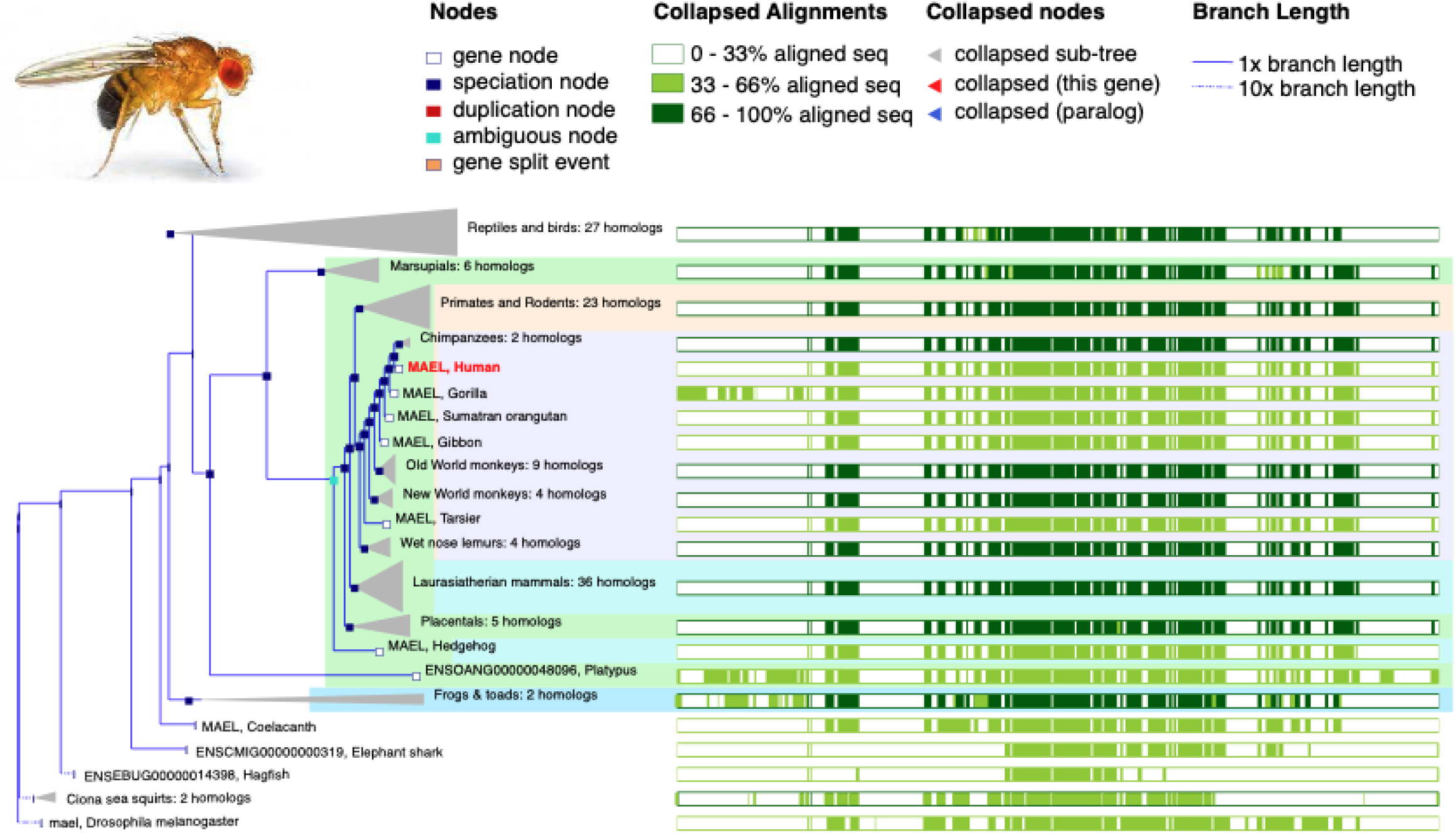
Phylogenetic tree demonstrating the evolutionary origin of *maelstrom* (*MAEL*), first arising in *Drosophila melanogaster*. Taxonomic analysis and visualization were performed using Ensembl. Gene tree analysis reveals evolutionary origin of MAEL across species using Ensembl ^16^. There are 127 total speciation nodes and 2 duplication events. Gene/loss family evolution overtime was performed as previously described ^17^. Figure created using Ensembl.

### Single cell RNA sequencing implicates copy number variations (CNVs) during prenatal human neurodevelopment

Since *MAEL* has been demonstrated to regulate DNA structure ^36^ and transposon regulation ^37^, which has been shown to induce large-scale genomic rearrangements including copy number variations (CNVs) in the human brain ^38–40^, we first wanted to investigate the landscape of somatic CNVs during prenatal human brain development. In addition, it is estimated that 200-400 somatic single nucleotide variants (SNVs) are present in the brain at mid-gestation ^41^. Thus, we aimed to quantify the somatic CNV burden in the prenatal brain across time points. While scRNA-seq is traditionally employed to profile transcriptomic states at single-cell resolution, it can also be leveraged to infer large-scale CNVs through expression-based approaches. Thus, we analyzed single nuclei RNA sequencing (snRNA-seq) data from over 200,000 nuclei derived from the proliferative germinal matrix and laminating cortical plates of prenatal, non-pathological postmortem samples from 17 to 41 gestational weeks (GW) ^19^. We applied inferCNV ^42^ to four samples (one in each prenatal stage) from the single cell data. As shown in Figure 2, the CNV diversity in each sample from each prenatal stage is marked, with a cell-type specific preference and even within a cell type. This suggests that somatic CNVs are likely to exist in the prenatal brain tissue. Thus we hypothesize that DNA transposons and epigenetic regulation during this critical period of large-scale genomic rearrangement that dictates cell identity and lineage is likely to be altered by *MAEL* expression. However, the specific cell types and direction of *MAEL* expression in the neonatal (HC) human brain is unknown.

**Figure 2.**
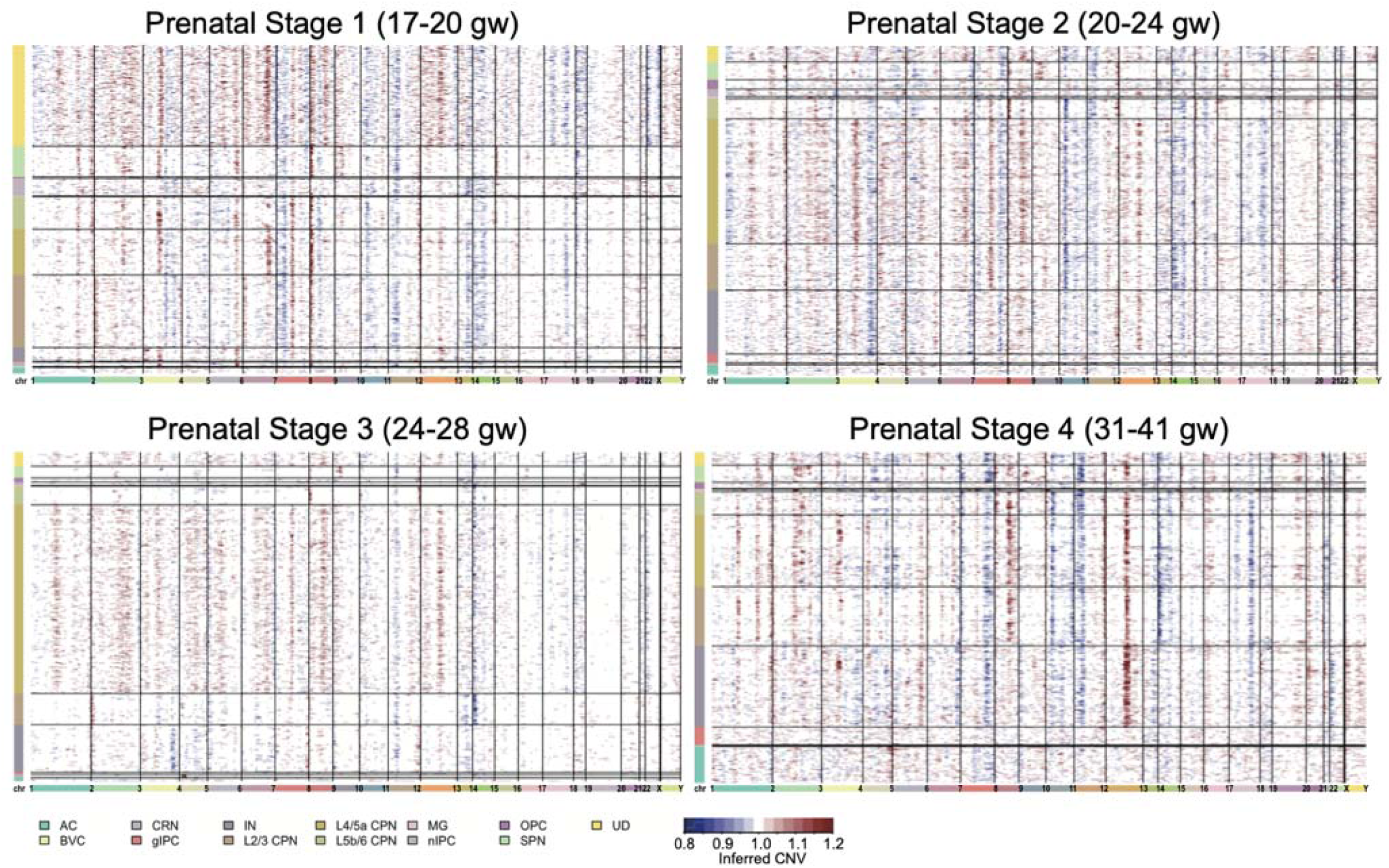
Single cell data revealed somatic copy number variations (CNVs) in prenatal brain. Four samples (sample IDs: 24, 34, 62, 6) from four prenatal stages are shown ^53^. Chromosomes are shown in the x-axis. Cell types are shown in the y-axis. Abbreviations are as follows: AC astrocyte, BVC blood vessel cell, CRN Cajal Retzius Cell, gIPC glial intermediate progenitor cell, IN interneuron, CPN cortical projection neuron, L2/3, L4, L5/6 neocortical layers 2/3, 4, 5/6, MG microglia, nIPC neuronal intermediate progenitor cell, OPC oligodendrocyte progenitor cell, SPN subplate neuron, UD not included, gw gestational weeks.

### Single-cell RNA sequencing of the neonatal brain across developmental timescales reveals cellular origin of MAEL

The emergence of large-scale sequencing efforts and biorepositories (i.e., establishing ‘normal’) enables inference of gene-based disease mechanisms. Gene expression is a dynamic process as cells proliferate, develop, and form 3D structures. Thus, our aim was to identify the spatiotemporal landscape of *MAEL* expression in the developing human brain. The Allen Brain Atlas constructed a single-cell RNA sequencing database of neonatal brain tissue across developmental timescales ^21,43^. First, we considered MAEL expression across time and utilized the Developing Human Brain Atlas (Allen Institute), which contains 49 unique brain regions. We used principal component analysis (PCA) to delineate *MAEL* expression across 13-time points, including 7 prenatal, 2 infancy, 2 childhood, 1 adolescence, and 1 adult epochs (Figure 3). These data demonstrate approximately 3 unique expression clusters – early prenatal, late prenatal, and late childhood/adulthood. We hypothesize that aberrant *MAEL* regulation during these sensitive developmental windows, alone and in concert with a “second hit” event including intraventricular hemorrhage (IVH), infection, hypoxia etc., may trigger development of HC.

**Figure 3.**
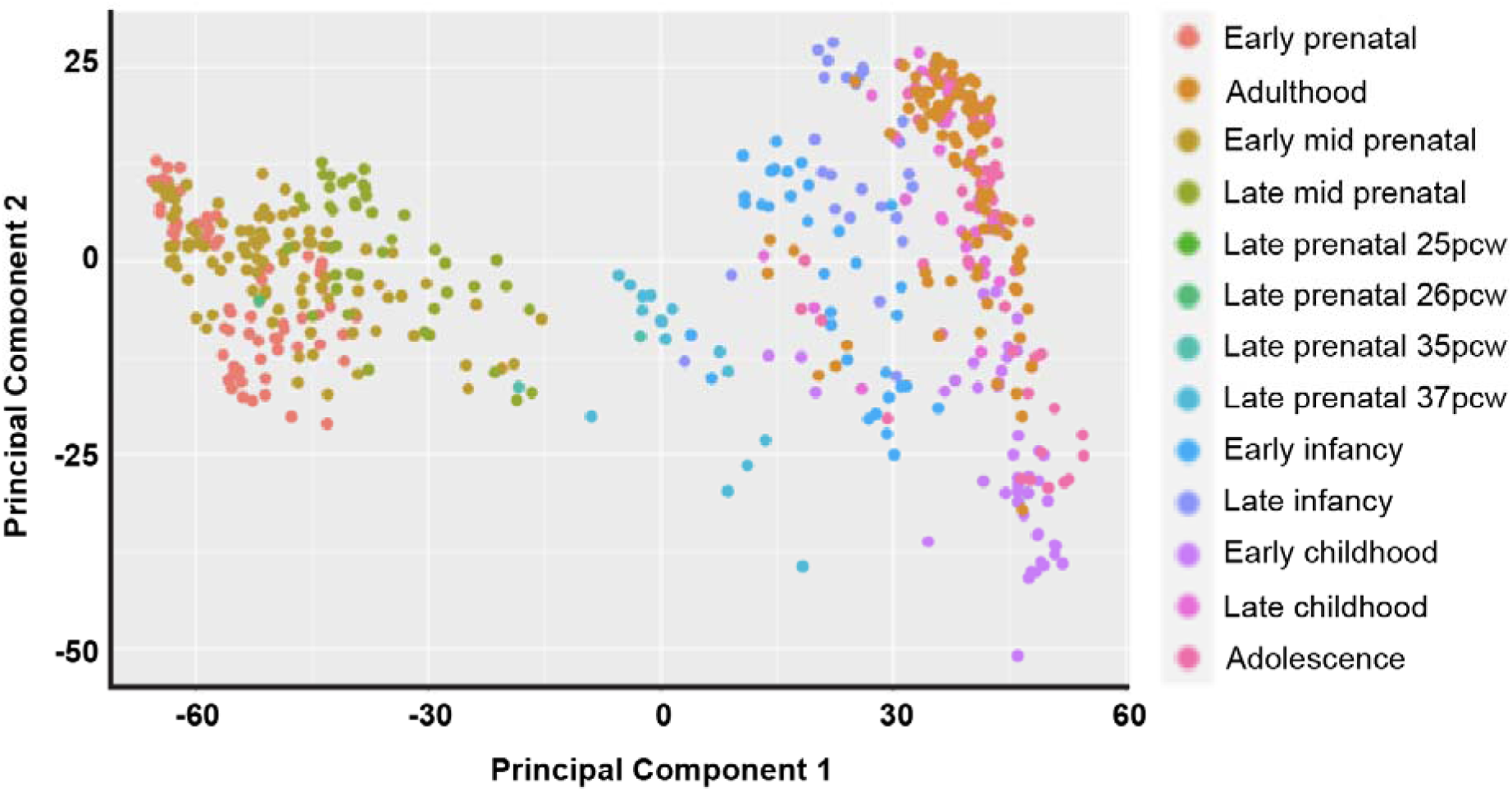
*MAEL* expression across timescales in the developing human brain. We analyzed the Developing Human Brain Atlas (Allen Institute) containing 49 brain regions at to identify temporal and spatial expression patterns. Principal components analysis (PCA) was performed across developmental time frames.

These data demonstrate that global *MAEL*-associated expression profiles segregate by developmental stage, indicating that expression dynamics vary across time (Figure 3). Next, our aim was to quantify *MAEL* expression over time and across embryologic origin in the developing brain. To improve statistical power and infer potential biologic mechanisms, we grouped brain regions into telencephalon, metencephalon, and diencephalon across the 13 epochs described above (Figure 4). *MAEL* expression in the telencephalon (cortex, forebrain and hippocampus) is very low and then gradually increases over time, peaking in late childhood, whereas *MAEL* expression is highest in the metancephalon (cerebellum and pons) during development and then decreases over time. Finally, *MAEL* expression in the diencephalon (thalamus, hypothalamus, and posterior pituitary) is relatively static. These data suggest that if the genetic component of MAEL expression restricts its expression across developmental, the “brake” on transposon activity is lost leading to aberrant cell identity and function. More broadly, these data highlight the necessity of accounting for time and tissue type when performing gene-based association analyses to guide hypothesis-driven experiments.

**Figure 4.**
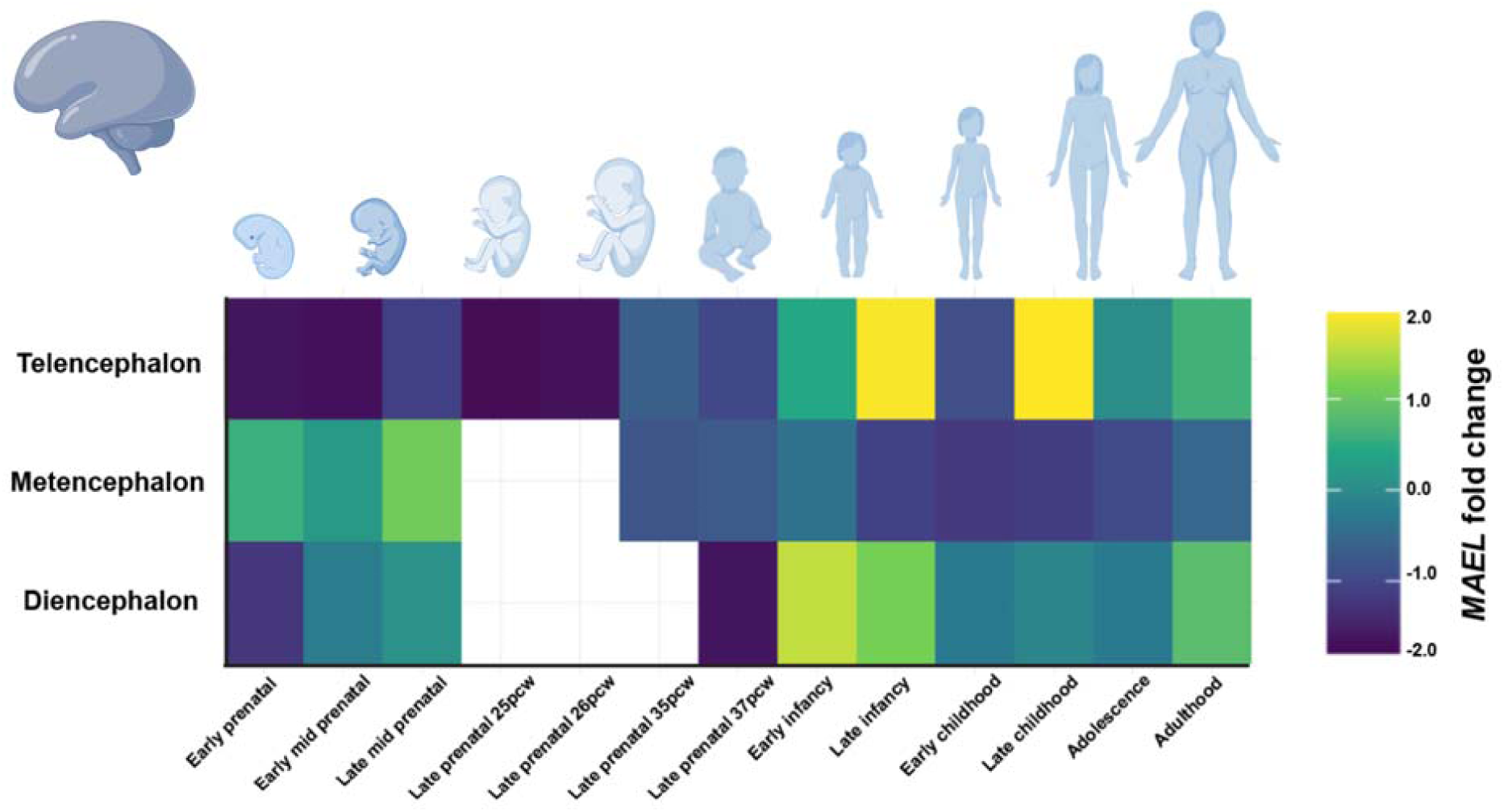
*MAEL* expression across time and embryologic origin. We analyzed the Developing Human Brain Atlas (Allen Institute) containing 49 brain regions, grouped by embryologic origin, to improve statistical power and infer how impaired *MAEL* expression may contribute to hydrocephalus risk at different ages. Since MAEL expression varies considerable across tissues and age, considering time as a variable in gene-based association analyses may be highly relevant in genetic studies of hydrocephalus.

In addition, we analyzed data across three developmental time points (15, 16, and 21 post-conception weeks (pcw)). These data revealed *MAEL* expression lowest in the inner subventricular zone (SVZ), inner SVZ ventrolateral prefrontal cortex, and outer SVZ posteroinferior parietal cortex at 16, 21, and 15 pcw, respectively. *MAEL* expression was highest in the outer SVZ caudal mid-inferior temporal cortex, inner SVZ posterior parahippocampal cortex, and outer SVZ lateral temporal-occipital cortex at 16, 16, and 21 pcw, respectively (Table 1). These data suggest potential regions and timepoints where different expression of *MAEL* may confer risk to hydrocephalus.

**Table 1.** Single-cell RNA sequencing across space and time reveals developmental origin of *MAEL* in the neonatal human brain. The Allen Brain Atlas Developing Brain resource ^21^was used to query MAEL expression across three developmental time points (15, 16, and 21 post-conception weeks (pcw)) from two donors. SZ is abbreviation of subventricular zone.

| Donor ID | Donor age | Structure Name | Donor 1 | Donor 2 | average |
| --- | --- | --- | --- | --- | --- |
| 14751 | 16 pcw | inner SZ in primary visual cortex | -3.32 | -2.57 | -2.95 |
| 12690 | 21 pcw | inner SZ in ventrolateral prefrontal cortex | -2.85 | -2.62 | -2.73 |
| 12840 | 15 pcw | outer SZ in posteroinferior (ventral) parietal cortex | -2.64 | -1.64 | -2.14 |
| 12840 | 15 pcw | inner SZ in dorsomedial frontal cortex | -2.60 | -1.59 | -2.09 |
| 12840 | 15 pcw | outer plexiform zone in extrastriate cortex | -2.29 | -1.49 | -1.89 |
| 12840 | 15 pcw | inner SZ in midlateral temporal cortex | -2.18 | -1.51 | -1.85 |
| 14751 | 16 pcw | inner SZ in posterior frontal cortex (motor cortex) | -1.28 | -2.30 | -1.79 |
| 12566 | 21 pcw | inner SZ in ventrolateral prefrontal cortex | -1.04 | -2.44 | -1.74 |
| 14751 | 16 pcw | outer SZ in frontal polar cortex | -1.59 | -1.73 | -1.66 |
| 12840 | 15 pcw | outer SZ in primary somatosensory cortex | -1.32 | -1.97 | -1.65 |
| 14751 | 16 pcw | outer SZ in posterior parahippocampal cortex | -0.97 | -2.16 | -1.56 |
| 12840 | 15 pcw | inner SZ in posterior frontal cortex (motor cortex) | -1.67 | -1.34 | -1.51 |
| 12840 | 15 pcw | SZ in caudal cingulate cortex | -1.23 | -1.74 | -1.48 |
| 12840 | 15 pcw | outer SZ in orbital frontal cortex | -1.52 | -1.39 | -1.45 |
| 12840 | 15 pcw | inner SZ in inferolateral temporal cortex | -1.51 | -1.39 | -1.45 |
| 12840 | 15 pcw | inner SZ in dorsolateral prefrontal cortex | -0.90 | -1.95 | -1.42 |
| 14751 | 16 pcw | inner SZ in primary somatosensory cortex | -0.57 | -2.17 | -1.37 |
| 14751 | 16 pcw | inner SZ in medial temporal-occipital cortex | -0.87 | -1.87 | -1.37 |
| 12566 | 21 pcw | inner SZ in primary visual cortex | -1.11 | -1.58 | -1.35 |
| 12840 | 15 pcw | inner SZ in dorsomedial parietal cortex (area 7m) | -1.13 | -1.42 | -1.27 |
| 12690 | 21 pcw | inner SZ in posterior frontal cortex (motor cortex) | -0.91 | -1.59 | -1.25 |
| 14751 | 16 pcw | inner SZ in posteroinferior (ventral) parietal cortex | -0.67 | -1.78 | -1.23 |
| 12840 | 15 pcw | outer SZ in midlateral temporal cortex | -0.67 | -1.63 | -1.15 |
| 12690 | 21 pcw | outer SZ in inferolateral temporal cortex | -0.78 | -1.41 | -1.10 |
| 12690 | 21 pcw | outer SZ in posterior frontal cortex (motor cortex) | -1.18 | -0.91 | -1.04 |
| 12840 | 15 pcw | inner SZ in posterosuperior (dorsal) parietal cortex | -0.90 | -1.08 | -0.99 |
| 12840 | 15 pcw | outer SZ in dorsomedial frontal cortex | -0.60 | -1.39 | -0.99 |
| 12566 | 21 pcw | SZ in caudal cingulate cortex | -0.81 | -1.16 | -0.98 |
| 12690 | 21 pcw | inner SZ in orbital frontal cortex | -0.99 | -0.96 | -0.97 |
| 12840 | 15 pcw | inner SZ in primary somatosensory cortex | -0.66 | -1.22 | -0.94 |
| 14751 | 16 pcw | inner SZ in frontal polar cortex | 0.15 | -1.97 | -0.91 |
| 14751 | 16 pcw | inner SZ in ventromedial extrastriate cortex | -0.33 | -1.45 | -0.89 |
| 12566 | 21 pcw | inner SZ in primary somatosensory cortex | -0.95 | -0.83 | -0.89 |
| 14751 | 16 pcw | outer SZ in posterior frontal cortex (motor cortex) | -0.59 | -1.18 | -0.88 |
| 12840 | 15 pcw | outer SZ in midlateral extrastriate cortex | -0.65 | -1.10 | -0.87 |
| 12690 | 21 pcw | inner SZ in posterosuperior (dorsal) parietal cortex | -0.97 | -0.76 | -0.87 |
| 12566 | 21 pcw | outer SZ in posterosuperior (dorsal) parietal cortex | -0.53 | -1.17 | -0.85 |
| 14751 | 16 pcw | inner SZ in dorsomedial parietal cortex (area 7m) | -0.75 | -0.92 | -0.84 |
| 12690 | 21 pcw | outer SZ in posterosuperior (dorsal) parietal cortex | -0.61 | -1.00 | -0.80 |
| 12690 | 21 pcw | outer SZ in posteroinferior (ventral) parietal cortex | -0.60 | -0.95 | -0.77 |
| 12690 | 21 pcw | inner SZ in dorsomedial frontal cortex | -0.56 | -0.96 | -0.76 |
| 12566 | 21 pcw | outer SZ in dorsomedial parietal cortex (area 7m) | -0.56 | -0.89 | -0.72 |
| 14751 | 16 pcw | outer plexiform zone in extrastriate cortex | -0.76 | -0.68 | -0.72 |
| 14751 | 16 pcw | outer SZ in posterosuperior (dorsal) parietal cortex | -0.66 | -0.78 | -0.72 |
| 12566 | 21 pcw | inner SZ in inferolateral temporal cortex | -0.81 | -0.60 | -0.70 |
| 14751 | 16 pcw | SZ in midcingulate cortex | -0.77 | -0.63 | -0.70 |
| 14751 | 16 pcw | inner SZ in dorsolateral prefrontal cortex | -0.51 | -0.88 | -0.70 |
| 14751 | 16 pcw | outer SZ in posteroinferior (ventral) parietal cortex | -0.62 | -0.77 | -0.69 |
| 12840 | 15 pcw | outer SZ in dorsomedial parietal cortex (area 7m) | -0.78 | -0.59 | -0.69 |
| 12840 | 15 pcw | SZ in subgenual cingulate cortex | -0.41 | -0.97 | -0.69 |
| 12566 | 21 pcw | outer SZ in ventromedial extrastriate cortex | -0.78 | -0.57 | -0.67 |
| 14751 | 16 pcw | SZ in caudal cingulate cortex | -0.42 | -0.91 | -0.67 |
| 12840 | 15 pcw | inner SZ in midlateral extrastriate cortex | -0.62 | -0.71 | -0.66 |
| 12840 | 15 pcw | inner SZ in lateral temporal-occipital cortex | -0.63 | -0.67 | -0.65 |
| 12690 | 21 pcw | outer SZ in ventrolateral prefrontal cortex | -0.19 | -1.09 | -0.64 |
| 12566 | 21 pcw | inner SZ in posteror frontal cortex (motor cortex) | -0.78 | -0.50 | -0.64 |
| 12566 | 21 pcw | outer SZ in primary visual cortex | -0.44 | -0.80 | -0.62 |
| 12690 | 21 pcw | inner SZ in inferolateral temporal cortex | -0.74 | -0.48 | -0.61 |
| 14751 | 16 pcw | inner SZ in posterosuperior (dorsal) parietal cortex | -0.35 | -0.86 | -0.61 |
| 12840 | 15 pcw | outer SZ in frontal polar cortex | -0.37 | -0.81 | -0.59 |
| 12840 | 15 pcw | outer SZ in posteror frontal cortex (motor cortex) | -0.35 | -0.80 | -0.58 |
| 12566 | 21 pcw | inner SZ in ventromedial extrastriate cortex | -0.73 | -0.42 | -0.58 |
| 14751 | 16 pcw | outer SZ in ventromedial extrastriate cortex | -0.57 | -0.58 | -0.58 |
| 12840 | 15 pcw | SZ in rostral cingulate cortex | -0.48 | -0.65 | -0.56 |
| 12690 | 21 pcw | inner SZ in primary somatosensory cortex | -0.79 | -0.31 | -0.55 |
| 12566 | 21 pcw | inner SZ in frontal polar cortex | -0.66 | -0.44 | -0.55 |
| 12690 | 21 pcw | outer SZ in dorsomedial frontal cortex | -0.71 | -0.35 | -0.53 |
| 12566 | 21 pcw | outer SZ in midlateral extrastriate cortex | -0.55 | -0.51 | -0.53 |
| 12566 | 21 pcw | inner SZ in dorsolateral prefrontal cortex | -0.54 | -0.47 | -0.50 |
| 14751 | 16 pcw | SZ in rostral cingulate cortex | -0.16 | -0.85 | -0.50 |
| 12840 | 15 pcw | outer SZ in ventrolateral prefrontal cortex | -0.62 | -0.37 | -0.49 |
| 12566 | 21 pcw | outer SZ in ventrolateral prefrontal cortex | -0.41 | -0.56 | -0.49 |
| 12840 | 15 pcw | outer SZ in ventromedial extrastriate cortex | -0.86 | -0.12 | -0.49 |
| 12566 | 21 pcw | outer SZ in primary somatosensory cortex | -0.45 | -0.51 | -0.48 |
| 12566 | 21 pcw | inner SZ in posteroinferior (ventral) parietal cortex | -0.55 | -0.39 | -0.47 |
| 12840 | 15 pcw | outer SZ in lateral temporal-occipital cortex | -0.38 | -0.56 | -0.47 |
| 12566 | 21 pcw | inner SZ in posterosuperior (dorsal) parietal cortex | -0.51 | -0.41 | -0.46 |
| 12690 | 21 pcw | inner SZ in midlateral extrastriate cortex | -0.46 | -0.46 | -0.46 |
| 12690 | 21 pcw | outer SZ in dorsolateral prefrontal cortex | -0.34 | -0.58 | -0.46 |
| 12840 | 15 pcw | inner SZ in medial temporal-occipital cortex | -0.45 | -0.44 | -0.45 |
| 14751 | 16 pcw | inner SZ in inferolateral temporal cortex | -0.41 | -0.43 | -0.42 |
| 12566 | 21 pcw | SZ in posterior parahippocampal cortex | -0.35 | -0.47 | -0.41 |
| 12840 | 15 pcw | inner SZ in dorsomedial extrastriate cortex | -0.31 | -0.47 | -0.39 |
| 12840 | 15 pcw | inner SZ in posteroinferior (ventral) parietal cortex | -0.31 | -0.44 | -0.37 |
| 12840 | 15 pcw | inner SZ in ventromedial extrastriate cortex | -0.22 | -0.51 | -0.37 |
| 12566 | 21 pcw | outer SZ in frontal polar cortex | -0.46 | -0.26 | -0.36 |
| 12690 | 21 pcw | outer SZ in midlateral extrastriate cortex | -0.35 | -0.35 | -0.35 |
| 12840 | 15 pcw | outer SZ in superolateral temporal cortex | -0.40 | -0.27 | -0.34 |
| 12566 | 21 pcw | outer SZ in dorsomedial extrastriate cortex | -0.44 | -0.20 | -0.32 |
| 14751 | 16 pcw | inner SZ in dorsomedial extrastriate cortex | -0.36 | -0.23 | -0.30 |
| 12840 | 15 pcw | inner SZ in primary visual cortex | -0.18 | -0.40 | -0.29 |
| 14751 | 16 pcw | inner SZ in caudal midinferior temporal cortex (area TF) | -0.26 | -0.30 | -0.28 |
| 12690 | 21 pcw | inner SZ in posteroinferior (ventral) parietal cortex | -0.44 | -0.11 | -0.27 |
| 12840 | 15 pcw | outer SZ in posterosuperior (dorsal) parietal cortex | -0.59 | 0.06 | -0.27 |
| 12566 | 21 pcw | inner SZ in orbital frontal cortex | -0.27 | -0.20 | -0.23 |
| 12566 | 21 pcw | outer SZ in dorsolateral prefrontal cortex | -0.06 | -0.40 | -0.23 |
| 12690 | 21 pcw | inner SZ in dorsolateral prefrontal cortex | -0.17 | -0.28 | -0.23 |
| 12840 | 15 pcw | inner SZ in frontal polar cortex | -0.36 | -0.09 | -0.22 |
| 12566 | 21 pcw | inner SZ in midlateral extrastriate cortex | -0.18 | -0.25 | -0.22 |
| 14751 | 16 pcw | outer SZ in primary somatosensory cortex | -0.33 | -0.09 | -0.21 |
| 12566 | 21 pcw | SZ in midcingulate cortex | -0.31 | -0.03 | -0.17 |
| 12566 | 21 pcw | outer SZ in inferolateral temporal cortex | -0.15 | -0.18 | -0.17 |
| 14751 | 16 pcw | outer SZ in inferolateral temporal cortex | -0.19 | -0.08 | -0.14 |
| 12566 | 21 pcw | outer SZ in posteroinferior (ventral) parietal cortex | -0.39 | 0.14 | -0.12 |
| 14751 | 16 pcw | outer SZ in dorsomedial parietal cortex (area 7m) | -0.20 | -0.04 | -0.12 |
| 12690 | 21 pcw | inner SZ in frontal polar cortex | -0.31 | 0.08 | -0.12 |
| 12840 | 15 pcw | inner SZ in ventrolateral prefrontal cortex | -0.23 | 0.01 | -0.11 |
| 12566 | 21 pcw | outer SZ in posterior frontal cortex (motor cortex) | -0.11 | -0.09 | -0.10 |
| 12566 | 21 pcw | SZ in medial temporal-occipital cortex | -0.07 | -0.12 | -0.09 |
| 12566 | 21 pcw | inner SZ in dorsomedial extrastriate cortex | -0.21 | 0.02 | -0.09 |
| 12690 | 21 pcw | outer SZ in frontal polar cortex | -0.12 | -0.01 | -0.06 |
| 12840 | 15 pcw | outer SZ in dorsomedial extrastriate cortex | 0.20 | -0.31 | -0.06 |
| 14751 | 16 pcw | outer SZ in superolateral temporal cortex | -0.22 | 0.21 | 0.00 |
| 12566 | 21 pcw | inner SZ in lateral temporal-occipital cortex | -0.07 | 0.08 | 0.00 |
| 14751 | 16 pcw | outer SZ in dorsomedial extrastriate cortex | -0.14 | 0.15 | 0.00 |
| 12840 | 15 pcw | outer SZ in dorsolateral prefrontal cortex | 0.14 | -0.11 | 0.01 |
| 12840 | 15 pcw | outer SZ in primary visual cortex | -0.09 | 0.13 | 0.02 |
| 12840 | 15 pcw | outer SZ in inferolateral temporal cortex | 0.01 | 0.07 | 0.04 |
| 12690 | 21 pcw | outer SZ in primary somatosensory cortex | 0.05 | 0.06 | 0.05 |
| 12690 | 21 pcw | outer SZ in orbital frontal cortex | -0.10 | 0.21 | 0.06 |
| 14751 | 16 pcw | outer SZ in medial temporal-occipital cortex | 0.36 | -0.24 | 0.06 |
| 14751 | 16 pcw | inner SZ in orbital frontal cortex | 0.27 | -0.12 | 0.08 |
| 12840 | 15 pcw | inner SZ in orbital frontal cortex | -0.04 | 0.22 | 0.09 |
| 12840 | 15 pcw | outer SZ in medial temporal-occipital cortex | -0.07 | 0.27 | 0.10 |
| 14751 | 16 pcw | SZ in subgenual cingulate cortex | 0.16 | 0.10 | 0.13 |
| 12690 | 21 pcw | SZ in rostral cingulate cortex | 0.10 | 0.22 | 0.16 |
| 12566 | 21 pcw | SZ in rostral cingulate cortex | 0.27 | 0.17 | 0.22 |
| 14751 | 16 pcw | outer SZ in primary visual cortex | 0.19 | 0.33 | 0.26 |
| 12840 | 15 pcw | inner SZ in superolateral temporal cortex | 0.22 | 0.34 | 0.28 |
| 12690 | 21 pcw | SZ in midcingulate cortex | 0.20 | 0.36 | 0.28 |
| 12566 | 21 pcw | outer SZ in orbital frontal cortex | 0.28 | 0.35 | 0.31 |
| 12690 | 21 pcw | SZ in caudal cingulate cortex | 0.34 | 0.38 | 0.36 |
| 14751 | 16 pcw | outer SZ in dorsolateral prefrontal cortex | 0.44 | 0.31 | 0.37 |
| 14751 | 16 pcw | outer SZ in orbital frontal cortex | 0.25 | 0.55 | 0.40 |
| 14751 | 16 pcw | outer SZ in caudal midinferior temporal cortex (area TF) | 0.26 | 0.69 | 0.48 |
| 14751 | 16 pcw | inner SZ in posterior parahippocampal cortex | 0.57 | 0.51 | 0.54 |
| 12566 | 21 pcw | outer SZ in lateral temporal-occipital cortex | 0.29 | 0.85 | 0.57 |

We often observe onset of congenital HC in the late prenatal developmental period. This timeframe corresponds with development of the cortical plate and germinal matrix, where the SVZ is the residence of neural precursor cells (NPCs) ^13,44,45^. Since *MAEL* expression in the SVZ, where transposon activity is the highest and where we observe variable expression of *MAEL* (Table 1), we aimed to further scrutinize the transcriptional patterns within this region. Recently, a single-cell atlas of the late prenatal germinal matrix and cortical plate has been reported ^19^. Here we report *MAEL* expression across in the developing germinal matrix which demonstrates *MAEL* expression largely restricted to glial precursor cells (GPC, Figure 5a). In addition, we observe *MAEL* expression in cortical plate in GPCs as well as in L2/3 and L4/5a excitatory neurons (EN, Figure 5b). These data imply that the genetic component of *MAEL* expression in GPCs, providing further support for the NPC model of HC^3,44,46^, and ENs may contribute to HC pathophysiology and potential comorbid phenotypes.

**Figure 5.**
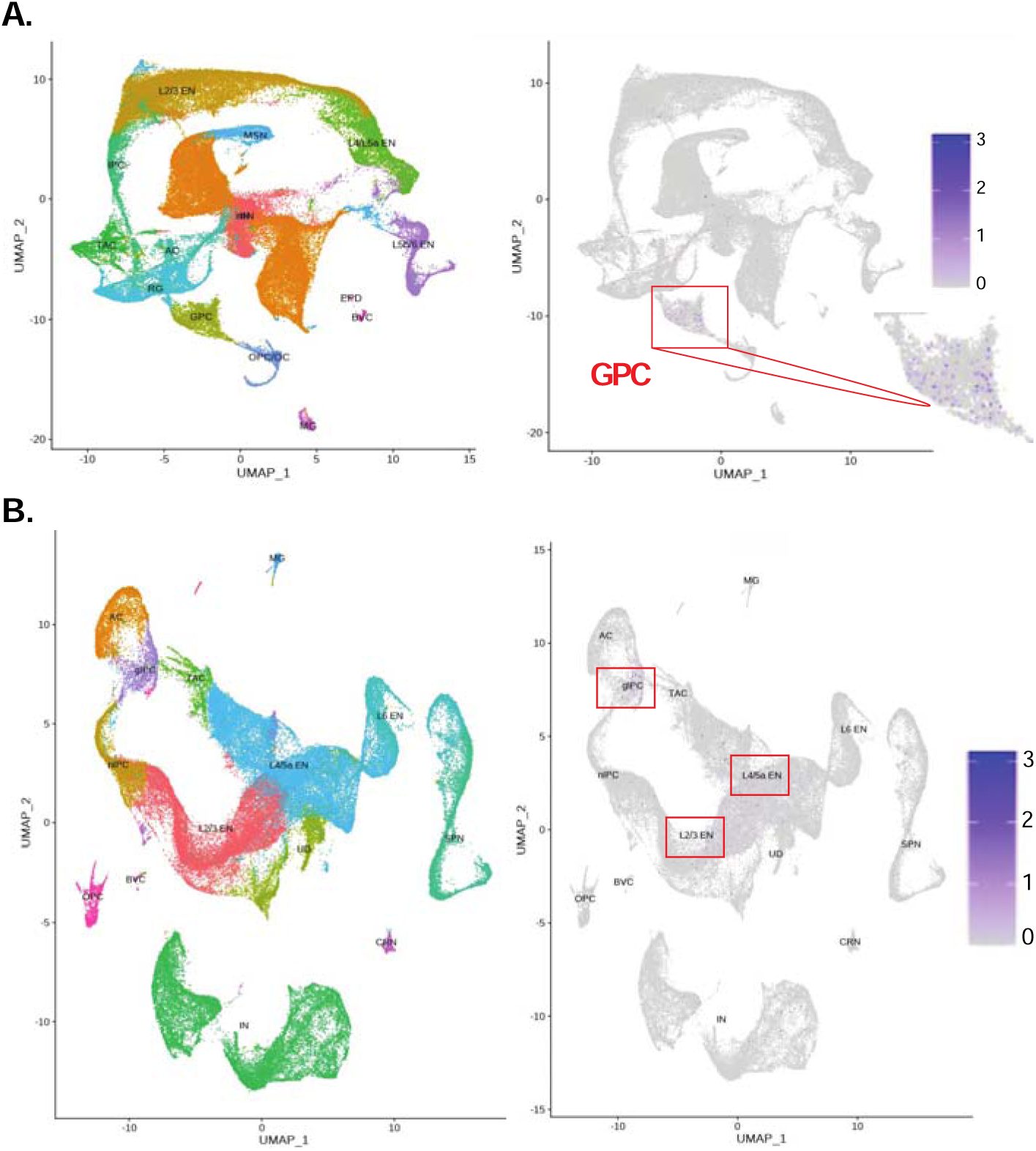
Single-cell RNA sequencing of *MAEL* in the developing germinal matrix (A) and cortical plate (B). We analyzed normal developing brain tissue as described in Ramos et al. *Nature Communications* (2022) ^19^. (A) *MAEL* expression is largely restricted to the glial precursor cell (GPC) population in the developing germinal matrix. (B) *MAEL* expression is largely restricted to GPCs, L2/3 excitatory neurons (EN), and L4/5a ENs in the developing cortical plate.

### Single-cell RNA sequencing brain of human hydrocephalus cortical brain tissue demonstrates reduced MAEL expression

Patients undergoing ETV were prospectively identified and consented for inclusion in our study (Table 2). Briefly, pre-frontal cortical tissue was obtained from the working channel of the endoscope and subjected to scRNA seq. To determine if directly measured *MAEL* expression was reduced in the human HC brain, consistent with prior findings from a human TWAS of HC ^14^, we quantified *MAEL* expression using single-nucleus RNA sequencing (snRNA-seq) data of human HC pre-frontal cortex. First, we quantified MAEL expression as a function of cell percentage within the ETV core sample.

**Table 2.**
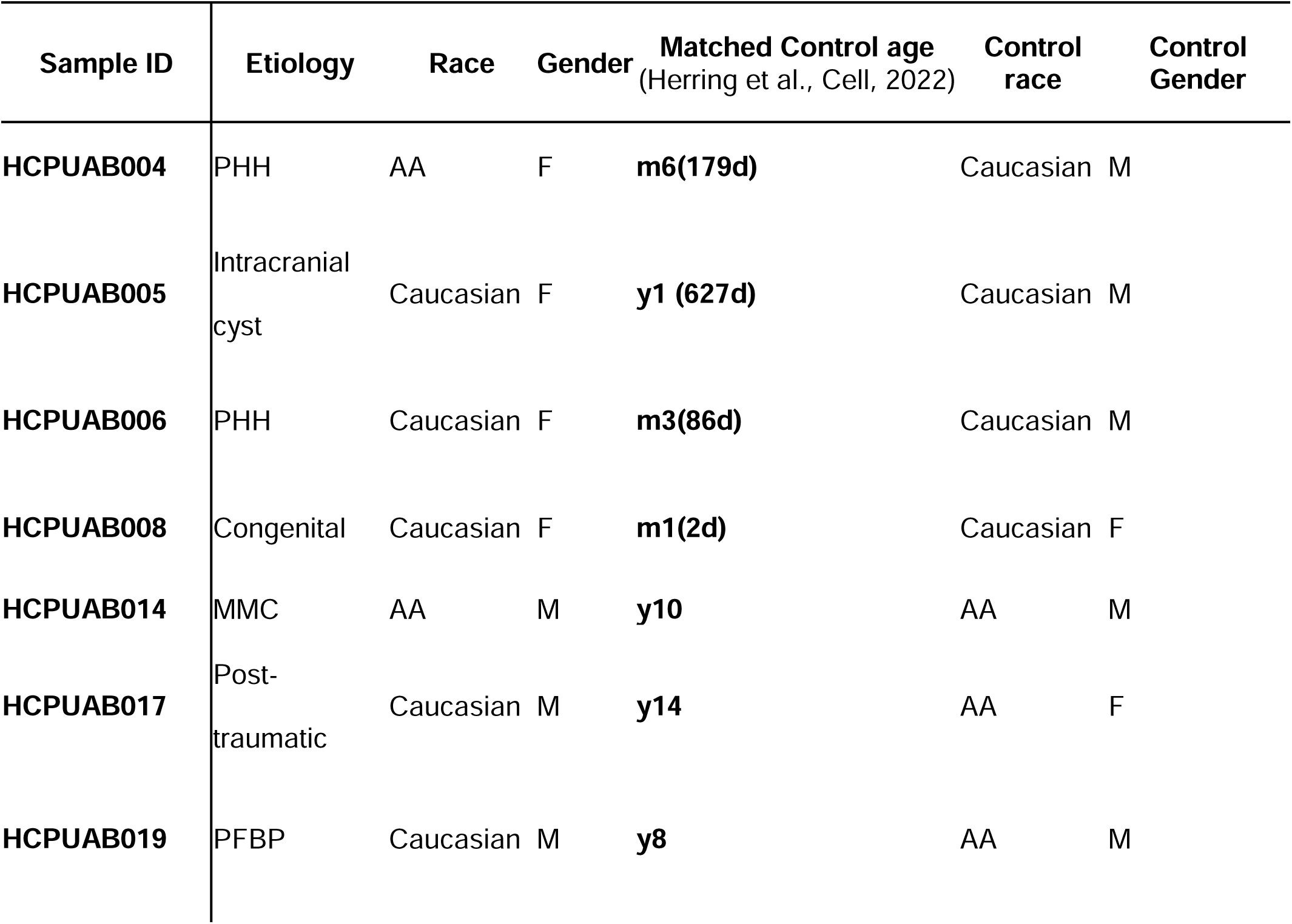
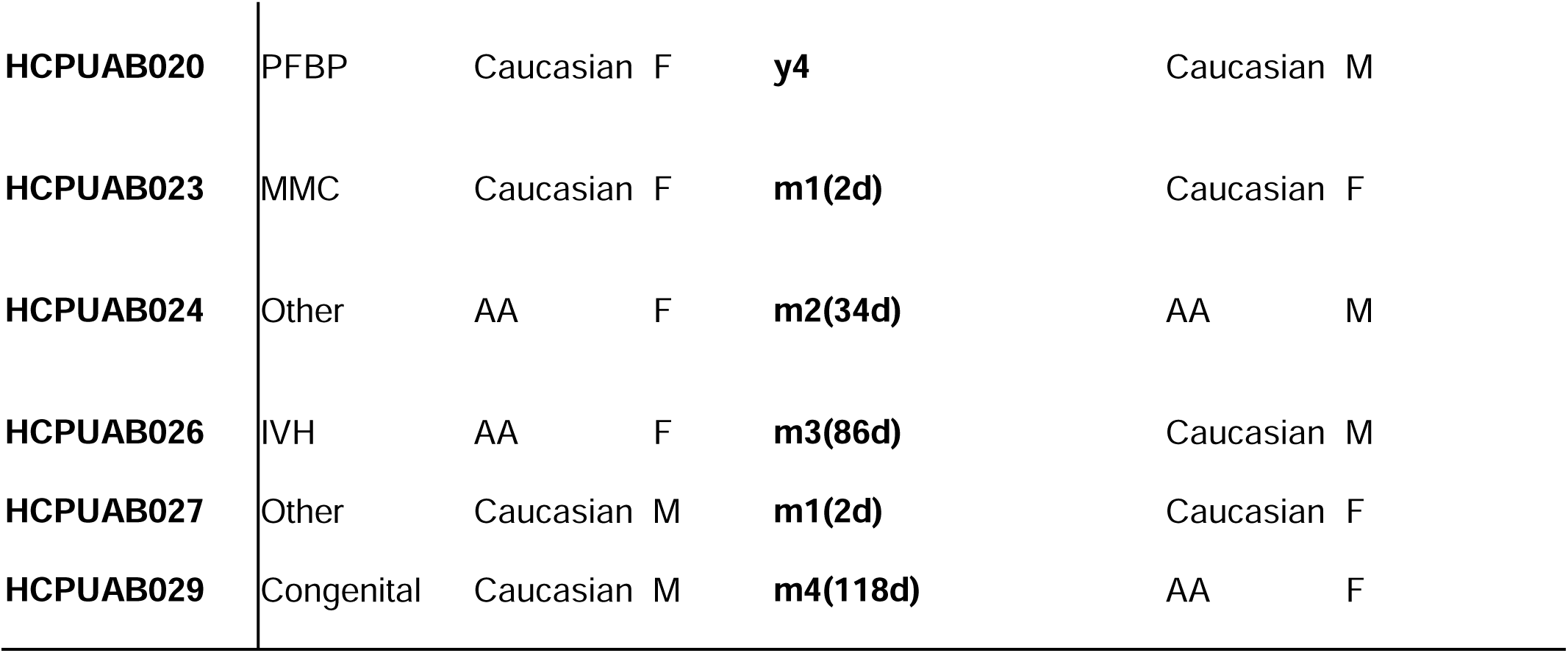
Patient characteristics of those undergoing ETV and subsequent scRNA sequencing. Matched control age and race from Herring et al. ^20^. Etiologies include post-hemorrhagic hydrocephalus (PHH), other intracranial cyst (e.g. arachnoid cyst, porencephalic), congenital communicating HC (congenital), myelomeningocele (MMC), posterior fossa brain tumor (PFBT), other (i.e., encephalocele), and post-traumatic HC. Race includes Caucasian and African American (AA) patients, as well as both female (F) and male (M).

We identified MAEL expression in < 10% of cells sampled and no clear expression pattern as a function of time in control samples (1 month – 14 years, Figure 6a). To recapitulate the gene-tissue pair TWAS analysis to differentiate between the genetic component and direct measurement of gene expression, we used pseudo-bulk analysis of scRNA-seq data and identified a statistically significant reduction in *MAEL* expression in the human HC brain (Figure 6b). However, because the majority of HC patients’ enrolled in our study are less than 3 years of age, this necessitated reference tissue atlases with improved resolution across this developmental time window. Thus, using age-matched experimental control non-diseased pre-frontal cortical tissue from Herring et al. ^20^, we aimed to quantify *MAEL* expression in individual cell types. Interestingly, we identified *MAEL* expression significantly reduced in most cell types (Figure 6c). These findings provide functional validation for reduced *MAEL* expression in HC pathogenesis ^14^. However, the precise molecular mechanisms by which reduced *MAEL* expression contributes to HC pathogenesis remains unknown.

**Figure 6.**
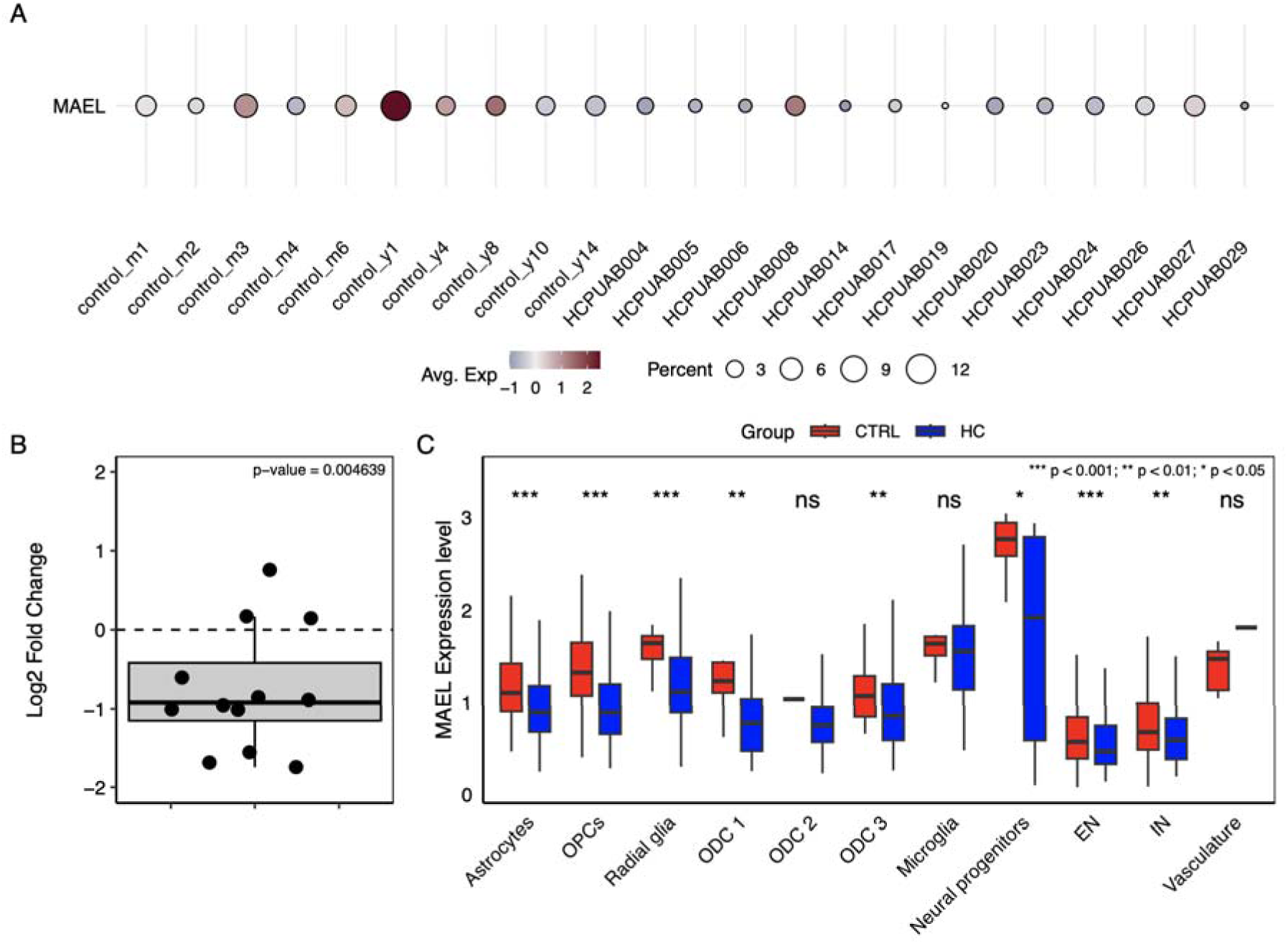
*MAEL* expression in the human hydrocephalic pre-frontal cortex. (A) Dot-plot demonstrating *MAEL* expression across cell percentage in individual human HC brain and age-matched controls as annotated in Herring et al ^20^. Dot size represents cell percentage and color represents expression level. (B) Psuedo-bulk analysis of *MAEL* expression in individual human HC pre-frontal cortical tissue. The central line represents the median, the box spans the interquartile range (IQR), and whiskers extend to 1.5×IQR from the hinges. A one-sample Wilcoxon signed-rank test was used. (C) Box and whisker plot demonstrating cell type-specific log_2_ fold change in *MAEL* expression compared between HC pre-frontal tissue and controls from Herring et al ^20^. Cells with zero expression were excluded, and group-wise differences were evaluated using the Wilcoxon rank-sum test with Benjamini–Hochberg correction. Outliers were suppressed for clarity, and adjusted p-values are indicated above each comparison. Statistical significance was reported as *** p < 0.001; ** p < 0.01; * p < 0.05.

## DISCUSSION

Here we systematically delineate the evolutionary, temporal, and spatial etiology of *MAEL* expression in the developing human brain using convergent and complementary integrative genomics approaches. We functionally validate *MAEL* expression in the human HC brain. This provides additional support for potential developmental, cell type, and temporal mechanisms for how *MAEL*, a transcriptome-wide predictor of HC ^14^, may contribute to HC risk. These data also implicated *MAEL* expression in ENs underlying HC, which may also contribute to comorbid neurodevelopmental disorders. Utilization of a human model system to elucidate HC molecular mechanisms will be required.

We provide multi-modal functional genomics data to elucidate the spatiotemporal expression patterns of *MAEL* in the developing human brain. We observed a substantial degree of CNV in the normal developing human brain (Figure 2), where MAEL may alter this process this process given its reported role in transposon and epigenetic regulation ^36,37^. However, the complete repertoire of cell-type specific transposon insertion sites in humans remains largely unknown ^47^. Furthermore, there is evidence that transposon elements are enriched in NPCs ^48^, where genetic variation or injury of this cell population contributes to select HC cases ^3,44,46^. We identify MAEL expression enriched in GPCs as well as ENs. To our knowledge, no patients with homozygous loss of function mutations in *MAEL*, have been identified, presumably owing to the necessity of this gene in development, utilization of human brain tissue and organoids may further clarify the mechanisms by which decreased *MAEL* causes large-scale genomic changes ^3^.

Many have demonstrated the role of gene expression and function of the ChP ^49^ and neural stem cell niche ^50,51^ play significant roles in HC pathogenesis. The relationship between neuronal and choroid plexus development, CSF dynamics, and genetic regulation underpin a highly-complex physiological system with multiple modes of regulation ^52^, converging on hallmark molecular mechanistic features ^3^. To understand the cellular origin and potential mechanistic impact of attenuated *MAEL* expression, we analyzed scRNA-seq data from the cortical plate and germinal matrix which harbor the developing NPCs ^19^. These data identified robust expression of *MAEL* in the neural stem cell compartment, where decreased MAEL expression may induce large-scale genomic rearrangements to influence cortical development, volume, and function. Delineating the impact of *MAEL* expression on brain growth and patterning during early embryogenesis may insights into HC pathobiology. Interestingly, we also identify MAEL highly expressed in ENs, which requires further functional validation for how aberrant development of these cells contribute to HC (and neurodevelopmental comorbidities).

We perform complementary and convergent functional genomics analysis delineating the origins of *MAEL* expression in non-disease primary human brain and HC brain tissue. These data provide the premise for mechanistic validation of this gene’s role in brain development and HC pathophysiology. Understanding genetic mechanisms of human disease necessitates human model systems, which in part, mirror the *in vivo* state. Transposon-mediated genetic regulation through *MAEL* expression is as a potential pathologic driver of HC, which may be directly tested in human brain organoid or assembloid systems. We perform evolutionary, temporal, and spatial single-cell transcriptomic analyses of the developing neonatal human brain and identify *MAEL* expression enriched in GPCs, L2/3 excitatory neurons (EN), and L4/5a ENs in the developing cortical plate. Finally, we provide direct functional evidence of reduced *MAEL* expression in the human HC brain. Collectively, our study extends observations made through integration of human genetics analysis with electronic health records, to functional genomics of, gene evolutionary, development, and *in vitro* human model systems.

## Data Availability

All data produced in the present study are available upon reasonable request to the authors.

## ACKNOWLEDGEMENTS

This work was supported by the National Institutes of Health (NIH) R21NS135321 (A.T.H. and Z.C.); Neurosurgery Research and Education Foundation (NREF) & American Academy of Neurological Surgery Grant, UAB Civitan Scholar Award, UAB Mary Heersink Institute for Global Health, Thrasher Early Career Award, Mission Brain Foundation Scholarship, NIH Fogarty Global Health Fellowship (Harvard University & University of Cape Town), and Oppenheimer Memorial Trust (A.T.H.); and R35GM138212 to Z.C.

## AUTHOR CONTRIBUTIONS

Conceptualization, A.T.H. and Z.C.; Methodology, A.T.H., Y.S., S.L., and Z.C.; Investigation, All authors; Writing – Original Draft, A.T.H.; Writing – Review and Editing, All Authors, Funding Acquisition – A.T.H., and Z.C., Supervision, A.T.H. and Z.C.

